# SARS-CoV-2 seroconversion in health care workers

**DOI:** 10.1101/2020.05.18.20105197

**Authors:** Adrian M. Shields, Sian E. Faustini, Marisol Perez-Toledo, Sian Jossi, Erin Aldera, Joel D. Allen, Saly Al-Taei, Claire Backhouse, Andrew Bosworth, Lyndsey Dunbar, Daniel Ebanks, Beena Emmanuel, Joanne Grey, I. Michael Kidd, Golaeh McGinnell, Dee McLoughlin, Gabriella Morley, Joanne O’Neill, Danai Papakonstantinou, Oliver Pickles, Charlotte Poxon, Megan Richter, Eloise Walker, Kasun Wanigasooriya, Yasunori Watanabe, Celina Whalley, Agnieszka E Zielinska, Max Crispin, David. C. Wraith, Andrew D. Beggs, Adam F. Cunningham, Mark. T. Drayson, Alex G. Richter

**Affiliations:** Clinical Immunology Service, Institute of Immunology and Immunotherapy, University of Birmingham, UK; University Hospital Birmingham NHS Foundation Trust, Birmingham, UK; Institute of Immunology and Immunotherapy, University of Birmingham, Birmingham, UK; Institute of Microbiology and Infection, University of Birmingham, Birmingham, UK; School of Biological Sciences, University of Southampton, Southampton, UK; PHE Public Health Laboratory, Birmingham, UK; Institute of Inflammation and Ageing, University of Birmingham, Birmingham, UK; Surgical Research Laboratory, Institute of Cancer and Genomics Science, University of Birmingham, UK; Oxford Glycobiology Institute, Department of Biochemistry, University of Oxford, Oxford, UK

## Abstract

**Background:** The correlates of protection against SARS-CoV-2 and their longevity remain unclear. Studies in severely ill individuals have identified robust cellular and humoral immune responses against the virus. Asymptomatic infection with SARS-CoV-2 has also been described, but it is unknown whether this is sufficient to produce antibody responses.

**Methods:** We performed a cross-sectional study recruiting 554 health care workers from University Hospitals Birmingham NHS Foundation Trust who were at work and asymptomatic. Participants were tested for current infection with SARS-CoV-2 by nasopharyngeal swab for real-time polymerase chain reaction and for seroconversion by the measurement of anti-SARS-CoV-2 spike glycoprotein antibodies by enzyme linked immunosorbent assay. Results were interpreted in the context of previous, self-reported symptoms of illness consistent with COVID-19.

**Results:** The point prevalence of infection with SARS-CoV-2, determined by the detection of SARS-CoV-2 RNA on nasopharnygeal swab was 2.39% (n=13/544). Serum was available on 516 participants. The overall rate of seroconversion in the cohort was 24.4% (n=126/516). Individuals who had previously experienced a symptomatic illness consistent with COVID-19 had significantly greater seroconversion rates than those who had remained asymptomatic (37.5% vs 17.1%, χ^2^ =21.1034, p<0.0001). In the week preceding peak COVID-19-related mortality at UHBFT, seroconversion rates amongst those who were suffering from symptomatic illnesses peaked at 77.8%. Prior symptomatic illness generated quantitatively higher antibody responses than asymptomatic seroconversion. Seroconversion rates were highest amongst those working in housekeeping (34.5%), acute medicine (33.3%) and general internal medicine (30.3%) with lower rates observed in participants working in intensive care (14.8%) and emergency medicine (13.3%).

**Conclusions:** In a large cross-sectional seroprevalence study of health-care workers, we demonstrate that asymptomatic seroconversion occurs, however prior symptomatic illness is associated with quantitatively higher antibody responses. The identification that the potential for seroconversion in health-care workers can associate differentially with certain hospital departments may inform future infection control and occupational health practices.

**Research in context:** *Evidence before the study:* To date, no study has examined the cross-sectional seroprevalence of anti-SARS-CoV-2 antibodies in health care workers during the COVID-19 pandemic. Existing evidence suggests that the levels of SARS-CoV-2 antibodies developing following infection may vary with disease severity in keeping with previous coronavirus pandemics.

*Added value of this study:* We demonstrate that seroconversion can occur in health care workers who have suffered no previous symptoms of SARS-Cov-2 infection. However, prior symptomatic infection tends to drive quantitatively superior antibody responses against the virus. We observed differential seroconversion rates in individuals working within different hospital departments. Using intensive care as a reference, the relative risk for seroconversion was greatest for those working in housekeeping, acute and general internal medicine.

*Implications of all the available evidence:* Insight into the current seroprevalence of SARS-CoV-2 antibodies within a high-risk cohort of health-care workers is of direct relevance as a reference point for future community serological surveys. We provide further evidence of asymptomatic infection and seroconversion, strengthening the argument for regular, routine screening of health-care workers. Finally, we provide evidence that individuals working in particular roles within the NHS are at greater risk of seroconversion with significant implications for their occupational health.

## Introduction

The correlates of protection against infection with SARS-CoV-2 and the immunological factors that determine progression to severe disease remain uncertain. Previous outbreaks of SARS provide limited insight as they focused on individuals with severe disease [1], a group that forms a small fraction of the overall number of COVID-19 cases [2]. Nevertheless, robust cellular and humoral responses are postulated to be necessary for temporary and certainly long-lasting immunity.

Individuals infected with SARS-CoV-2 requiring hospital admission have shown seroconversion rates approaching 100% two weeks after symptom onset [3]. Preliminary data suggests the magnitude of antibody responses against the virus and its neutralising capacity are proportional to age and strong correlations have been observed between the presence of nucleoprotein-specific T cells and neutralising antibody titres [4]. While asymptomatic detection of SARS-CoV-2 RNA has been demonstrated in health care workers and pregnant mothers [5, 6], it remains unclear whether asymptomatic infection can lead to an immune response, seroconversion and sterilising immunity.

The seroprevalence of anti-SARS-CoV-2 antibodies in health care workers and the general population is currently unknown. Cross-sectional studies that determine the seroprevalence of anti-SARS-CoV-2 antibodies can assist in defining prior exposure to the virus and, when followed longitudinally, inform the potential effectiveness of serum antibody status as a correlate of protection against future disease. Health care workers provide a unique group to understand the correlates of protection against SARS-CoV-2 because of their occupational exposure to the virus.

University Hospitals Birmingham NHS Foundation Trust (UHBFT) is one of the largest hospital trusts in the UK with over 20,000 employees delivering care to 2.2 million people per annum. We conducted a cross-sectional study of 554 staff at UHBFT to determine the incidence of infection and seroconversion in health care workers and their relationship to prior symptoms of COVID-19.

## Methods

A cross-sectional study of health-care workers at University Hospital Birmingham NHS Trust was undertaken, recruiting 554 individuals over the course of 24 hours on 25/4/2020 across four hospital sites serving central and east Birmingham. Invitation to participate in the study was made using UHBFT trust email. All individuals voluntarily provided a nasopharyngeal swab for SARS-CoV-2 polymerase chain reaction testing and blood and saliva samples for anti-SARS-CoV-2 testing on a high-sensitivity ELISA developed at the University of Birmingham. At enrolment, individuals were asked to retrospectively report any illnesses consistent with COVID-19 that they had suffered in the previous four months.

Serum was available for analysis on 516 individuals (**Table 1**). Detection of SARS-CoV-2 RNA was performed using real-time PCR (Viasure, CerTest Biotec) directed against the ORF1ab and N genes following guanidine isothiocyanate inactivation of nasopharyngeal swabs. Serological analysis was performed using a high-sensitivity ELISA developed in-house. Serological analysis was performed at biological containment level 2. High-binding plates (Greiner Bio-One) were coated with trimeric SARS-CoV-2 spike glycoprotein [7, 8] and blocked with Stabilcoat solution (Sigma Aldrich). Serum was pre-diluted 1:40 prior to analysis. A combined secondary layer containing horse-radish peroxidase conjugated polyclonal antibodies against IgG, IgA and IgM followed by 3,3′,5,5′-Tetramethylbenzidine development was used to detect the presence of antibodies. The cutoff for positivity on the ELISA was set at 2 standard deviations above the mean OD_450_ of eight pre-2019 negative sera run independently across seven separate plates.

**Table 1:** Demographics of study participants. Median and inter-quartile ranges are provided.

|  | All Participants | Seropositive | Seronegative | P |
| --- | --- | --- | --- | --- |
| **N** | 516 | 126 | 390 |  |
| **Age (years)** | 42<br>(30-51) | 41<br>(30-51) | 42<br>(31-51) |  |
| **Ethnicity** |  |  |  | 0.36 |
| - White (%) | 38.0 | 34.9 | 39.0 |  |
| - BAME (%) | 16.1 | 18.3 | 15.4 |  |
| - Not stated | 45.9 | 46.8 | 45.6 |  |
| **Gender** |  |  |  | 0.10 |
| - Male (%) | 24.8 | 19.0 | 26.7 |  |
| - Female (%) | 75.2 | 81.0 | 73.3 |  |

Patient mortality data for the UHBFT was sourced from data submitted to NHS England. Indices of deprivation in participants postcodes were sourced from 2019 national statistics published by the UK Ministry of Housing, Communities and Local Government [9].

Data were analysed using Graph Pad Prism 8.4.2 for macOS. Significant differences in categorical data were analysed using the Chi squared test. The Kolomogorov-Smirnov and Kruskal-Wallis tests were used to compare distributions of optical density data. The Mann-Whitney U test was used for all other purposes. Results were considered statistically significance if the p-value 0.05. The study was approved by the London - Camden & Kings Cross Research Ethics Committee, study number 282525. All participants provided written, informed consent prior to enrolment in the study.

## Results

The point prevalence of PCR positivity in asymptomatic health care workers at UHBFT on the 25/4/2020 was 2.39% (n=13/554).

Serum was available for further analysis on 516 individuals (**Table 1**). 26.3% (n=136/516) of these individuals reported a prior illness consistent with COVID-19. To examine the relationship between SARS-CoV-2 infection in staff and COVID-19 caseload throughout the trust, the onset of symptoms in health care workers with prior illnesses was mapped to weekly trust wide COVID-19 mortality and final seroconversion status on 25/4/2020 (**Figure 1a**). This showed that, in those experiencing prior illnesses, the highest rates of seroconversion (77.8%, n=14/18) were observed in the week beginning 28/3/2020, one week before the peak weekly mortality was reached within UHBFT. No one developed symptoms in the two weeks before 25/04/2020 because they were excluded from the study through self isolation at home. The overall seroconversion rate across the cohort was 24.4% (n=126/516); individuals reporting a prior symptomatic illness had significantly greater rates of seroconversion than those who had been asymptomatic throughout the time-period assessed (37.5% vs 17.1%, X^2^ =21.1034, p<0.0001) **(Figure 1b**). Furthermore, antibody responses in individuals who had experienced a prior symptomatic illness were quantitatively greater than those who remained asymptomatic (Kruskal-Wallis statistic 7.159, p=0.02) **(Figure 1c)**.

**Figure 1:**
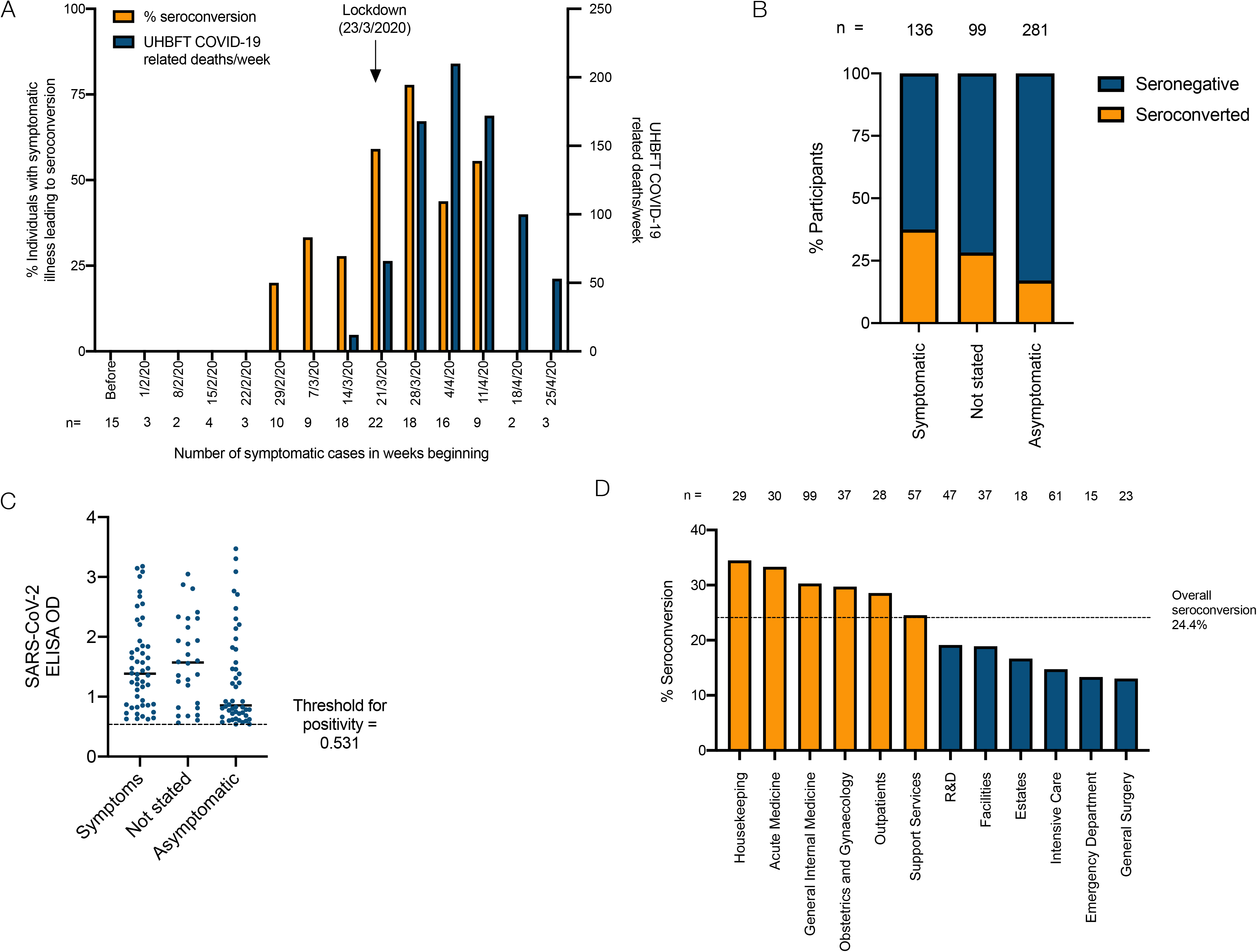
(A) Timing of prior symptomatic illness in study participants and their relationship with seroconversion and overall UHBFT-wide deaths in the weeks of March and April 2020. (B) Seroconversion rates in study participants self-reporting prior symptomatic illnesses consistent with COVID-19 compared to asymptomatic individuals. (C) Optical density of anti-SARS-CoV-2 ELISA individuals demonstrating seroconversion classified by self-reported prior symptomatic illness (n=126). (D) Seroconversion rates of study participants by department in which they work.

Rates of seroconversion were mapped to the departments where individuals work within UHBFT (**Figure 1d)**. Rates of seroconversion were highest in those working in housekeeping (34.5%, n=10/29), acute medicine (33.3%, n=10/30) and general internal medicine (30.3%, n=30/99) with the lowest rates observed in participants working in intensive care (14.8%, n=9/61) and emergency medicine (13.3%, n=2/15) and general surgery (13.0%, n=3/23). Using intensive care as a reference, the relative risk for seroconversion for those working in housekeeping was 2.34 (CI 1.03-5.36, p=0.04), acute medicine 2.25 (CI 1.03-4.97, p=0.04) and general internal medicine 2.05 (CI 1.05-4.03, p=0.04).

There was no significant difference in ethnodemographic data between individuals who had and had not seroconverted by the time of the study including indices of deprivation within the geographical postcode where study participants live (**Table 1 and Supplementary Figure 1)**. This supports an interpretation that the observed difference in seroconversion rates are more likely due to occupational risk, rather than external factors.

## Discussion

We present a cross-sectional study of health care workers that simultaneously determines the prevalence of infection and antibody seroconversion against SARS-CoV-2 in a group of healthcare workers employed by one of the largest NHS trusts in the United Kingdom.

The point prevalence of infection in health care workers in this study (2.39%) is concordant with the results of a similar study performed in London during the same week [5] and less than the rate of 14% observed in symptomatic health care workers in Newcastle on samples taken from March 10th to March 31st 2020 [10]. In contrast to these results, we report an overall seroconversion rate of 24.4% across our cohort by 25/4/2020. This would suggest that PCR testing on nasopharyngeal swabs consistently underestimates true infection rates [11]. In light of further evidence of asymptomatic infection and seroconversion, the impact of mandatory screening of health care workers should be thoroughly investigated [12]

The seroprevalence of SARS-CoV-2 antibodies amongst the UK general population remains unknown and few studies have considered seroprevalence in other populations. In Santa Clara, California, the seroprevalence rate has been estimated at between 1.3 and 4.7%, significantly lower than that found in the health-care worker cohort described herein, but this was assessed using different diagnostic methods [13]. In the SARS-CoV epidemic, the rate of seroconversion in healthcare workers with SARS (88.9%) and in asymptomatic health care workers (1.4%) were both higher than the general population (0.4%) suggesting increased seroprevalence due to occupational risk. This also indicates that, even after exposure to this far more lethal virus, seroconversion can occur without any accompanying symptomatology.

In our cohort, the relationship between prior symptomatic illness and seroconversion closely followed the temporal epidemiology of the pandemic across the UK and at UHBFT. We cannot exclude the possibility this study underestimates SARS-CoV-2 infections in health care workers by failing to capture more recent infections that had not led to seroconversion by 25th April. Longitudinal studies involving serial sampling will be necessary to provide up-dated assessments of seroconversion and the longevity of antibody responses.

Individuals with prior symptomatic illnesses were significantly more likely to seroconvert than asymptomatic individuals (35.8% vs 17.1%) and, in general, mount quantitatively greater antibody responses, in keeping with studies from the SARS-CoV pandemic [14]. Future work must explore the quality and longevity of antibody responses in individuals who have seroconverted, in particular, whether neutralising anti-SARS-CoV-2 antibodies persist and provide sterilising immunity [4].

Debate exists regarding nosocomial transmission of SARS-CoV-2 from patients to staff [10] [15]. Seroconversion rates in our cohort were highest amongst housekeepers and those working in acute and general internal medicine (32.7%). The explanation for this remains unclear; exposure to the virus is necessary for seroconversion and it is plausible that exposure to the virus is greater in these employees. In the 2003 SARS-CoV epidemic, seroconversion rates were highest amongst nurses and health-care assistants raising the possibility of differential occupational exposure and risk [16].

To explore the possibility that occupational exposure to SARS-CoV-2 is important compared with exposure from outside of the health-care setting, indices of deprivation were considered in the home postcodes of those who seroconverted compared to those who had not and no differences were observed. Deprivation has been associated with an increased risk of death from COVID-19, although whether this is partly secondary to increased exposure to the virus is not known [17]. Knowledge of local and population rates of seroconversion will allow more accurate determination of the relative risk of working in different hospital environments and better inform infection control and prevention measures.

In conclusion, we provide evidence of SARS-CoV-2 seroconversion in health care workers with and without prior symptomatic illness. The magnitude of the antibody response is greater in those with prior illness. We observe that the risk of seroconversion appears variable depending on where individuals work within the hospital environment.

## Data Availability

The authors confirm that the data supporting the findings of this study are available within the article.

## Acknowledgments

The authors would like to acknowledge the staff of the Clinical Immunology Service who helped process the samples for PCR and serological testing, Dr. Margaret Goodall for her expertise in antibody production and assay development, Dr Jason McLellan for the expression plasmid for the SARS-CoV-2 glycoprotein. We would also like to acknowledge all the participants from UHBFT. Serological assay development was undertaken in collaboration with The Binding Site Group Ltd.

---

**Supplementary Figure 1:**
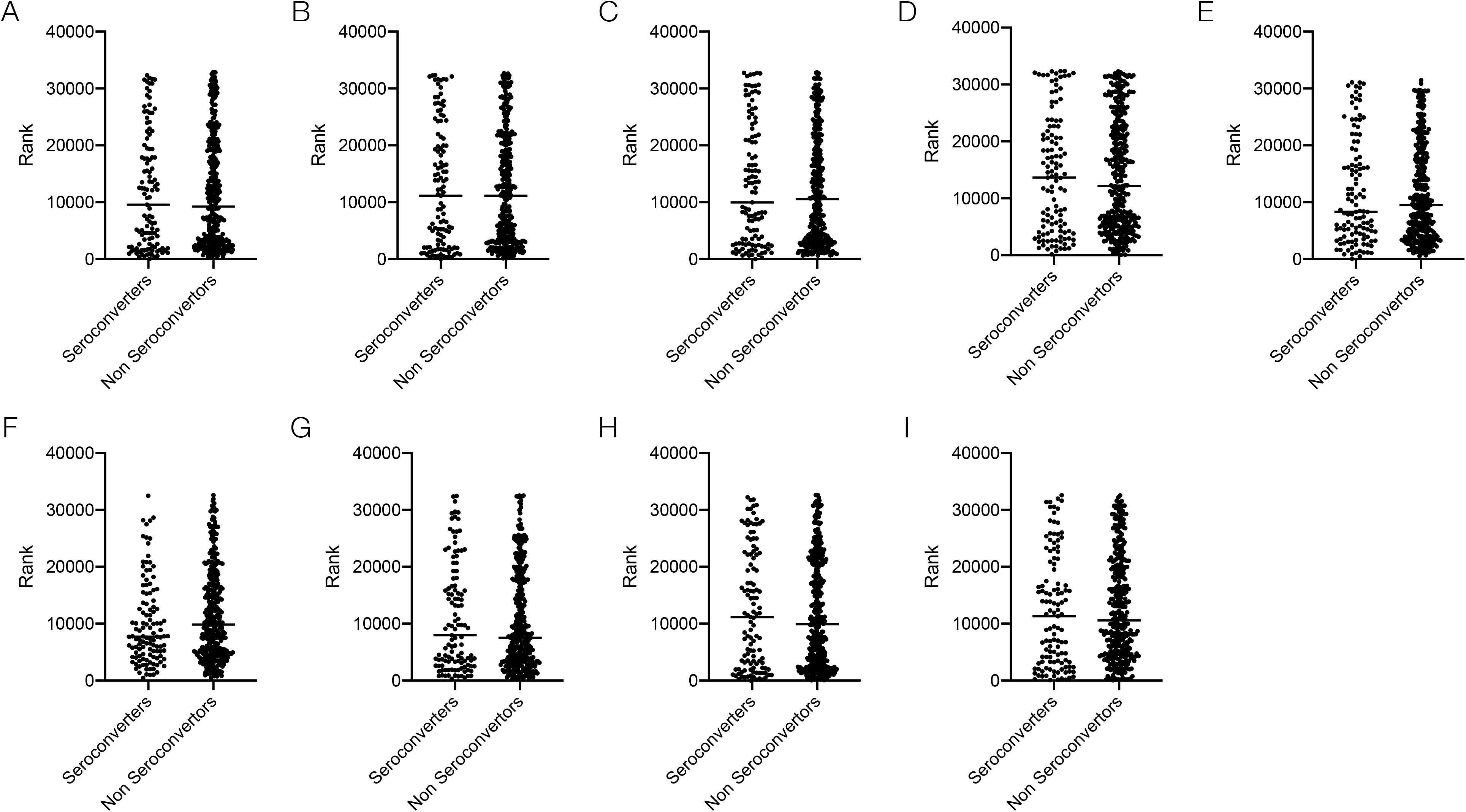
Indices on deprivation associated with home postcode of study participants: (A) Index of multiple deprivation, (B) Income rank, (C) employment rank, (D) education and skills rank, (E) Health and disability rank, (F) Barriers to housing and services rank, (G) Living environment rank, (H) Income deprivation affecting children index rank, (I) Income deprivation affecting adults rank.

